# Epigenome-wide association study of pregnancy exposure to green space and placental DNA methylation

**DOI:** 10.1101/2024.07.21.24310780

**Authors:** Sofía Aguilar-Lacasaña, Marta Cosin-Tomas, Bruno Raimbault, Laura Gómez, Olga Sánchez, Maria Julia Zanini, Rosalia Pascal Capdevila, Maria Foraster, Mireia Gascon, Ioar Rivas, Elisa Llurba, Maria Dolores Gómez-Roig, Jordi Sunyer, Martine Vrijheid, Mariona Bustamante, Payam Dadvand

**Author notes:** Joint last authors (Martine Vrijheid, Mariona Bustamante, Payam Dadvand).

## Abstract

Maternal exposure to green space during pregnancy has been associated with lower risk for adverse birth outcomes; however, the underlying biological mechanisms remain largely unknown. Epigenetic changes, such as DNA methylation (DNAm), might be one of the molecular mechanisms contributing to this association. The placenta is the key organ for foetal growth and development, but studies of prenatal green space exposure and placental DNAm are scarce and have not been conducted on a genome-wide scale. Here, we aimed to investigate the association between exposure to green space during pregnancy and epigenome-wide placental DNAm.

This study was conducted in 550 mother-child pairs from the Barcelona Life Study Cohort (BiSC), a population-based birth cohort in Spain (2018-2021). We comprehensively assessed green space exposure during pregnancy as (i) residential surrounding greenness (satellite-based Normalized Difference Vegetation Index (NDVI) in buffers of 100m, 300m and 500m), (ii) residential distance to the nearest major green space (meters), iii) use of green space (hours/week), and iv) visual access to greenery through home window (≥half of the view). Placental DNAm was measured with the EPIC array. To identify differentially methylated positions (DMPs), we fitted robust linear regression models adjusted for relevant covariates, while differentially methylated regions (DMRs) were identified using the *dmrff* method.

After Bonferroni (BN) correction, cg14852540, annotated to *SLC25A10* gene, was inversely associated with residential surrounding greenness within 500m buffer. Additionally, ten unique DMRs were associated with surrounding greenness within 300m and 500m, distance or visual access to green space. No associations were found for surrounding greenness within 100m or use of green space. Genes annotated to these loci are involved in transcriptional regulation, metabolism and mitochondrial respiration.

Overall, we identified associations between prenatal green space exposure and DNAm levels in placenta. Further research is needed to validate these results and understand the underlying biological pathways.

## 1. Introduction

Exposure to green space has been associated with health benefits over the entire life-course (Hu et al., 2021; Y. Yuan et al., 2020; Zare Sakhvidi et al., 2023). Specifically, prenatal exposure to green space has been associated with lower risk of adverse birth outcomes such as low birthweight (Nieuwenhuijsen et al., 2019; Torres Toda et al., 2022). Mechanistically, potential pathways underlying these associations include capacity instoration (e.g., encouraging physical activity and increasing social cohesion), harm mitigation (e.g., improving air quality, reducing noise, and dissipating heat, reducing exposure to air pollution, noise, and heat), capacity restoration (e.g., reducing stress and renewing attention) and immunity improvement (e.g., by enriching the microbiome) (Bowyer et al., 2022; Cruells et al., 2024; Markevych et al., 2017).

Epigenetic changes may be one of the molecular mechanisms contributing to the association between prenatal green space exposure and lower risk of adverse birth outcomes, either directly or indirectly through the aforementioned pathways. The epigenome comprises all modifications to DNA, or to DNA-associated RNA and proteins, that permit interpretation of the genome to instruct cell identity and function (Hemberger et al., 2020). These modifications ultimately influence gene expression. Among all epigenetic marks, DNA methylation (DNAm), the addition of a methyl group to the C5 position of the cytosine within a cytosine-guanine (CpG) dinucleotide, is the most widely investigated in epidemiological settings due to its relative stability with storage and the multitude of technical platforms available for analysis (Maccani & Marsit, 2009).

Recently, two genome-wide studies have explored the association between prenatal exposure to green space and DNAm in cord blood. One study, involving 538 neonates from Belgium, identified one significant CpG and 147 DMRs in association with exposure to green space (Alfano et al., 2023). The second study, involving 2,988 mother-infant pairs from seven European birth cohorts, found four DMRs associated with this exposure (Aguilar-Lacasaña et al., 2024). However, to better understand the intrauterine environment, it is crucial to investigate the placenta. This essential organ regulates nutrient and oxygen transfer and hormone and immune supply, and may act as a barrier to environmental exposures, playing a key role in foetal growth and development (Griffiths & Campbell, 2015; Mortillo & Marsit, 2022). To date, only a candidate gene study has explored the association between green space and placental DNAm, where the methylation status of the serotonin receptor *HTR2A* was positively associated with pregnancy green space exposure (Dockx et al., 2022). Thus, it is important to explore the association between maternal green space exposure during pregnancy and DNAm in placenta not only in specific genes but also on a genome-wide scale in relatively large samples sizes. Moreover, the aforementioned studies have mainly relied on residential surrounding greenness and/or proximity to a green space to assess green space exposure, overlooking other important aspects of this complex exposure such as use of green spaces or visual access to green space.

Our study aimed to fill these gaps by investigating the association between different aspects of maternal green space exposure during pregnancy and genome-wide placental DNAm. In addition to assessing residential greenness and proximity to green spaces, we also used data on the time spent in green spaces during pregnancy and the visual access to greenery through home windows.

## 2. Material and methods

### 2.1. Study population

The Barcelona Life Study Cohort (BiSC) is a prospective cohort study of 1,080 pregnant women, their offspring and partners in Barcelona that aims to identify early environmental and genetic causes of normal and abnormal growth, development and health from foetal life until young adulthood (Dadvand et al., 2024). Briefly, the enrolment of the BiSC participants was carried out between October 2018 and April 2021 at three tertiary university hospitals in Barcelona, Spain. Participants were recruited and had their first data collected at the end of the first trimester of pregnancy, with follow-ups in the second and third trimesters of pregnancy, delivery, and months 1, 2, 6, 8, 12, 18, 24, and 48 postnatally. Pregnant women aged 18-45 with singleton pregnancies were included. Exclusions applied to those residing outside the catchment area, not being able to communicate effectively in Spanish, Catalan or English, or with fetuses having known congenital anomalies. The study was approved by the ethical committees of the centers involved in the study, and written informed consent was obtained from all the participants. The detailed description of the recruitment of the study participants, data collection, and follow-ups are reported elsewhere (Dadvand et al., 2024). The total mother-child pairs with data on indicators of green space and placental DNAm included in this study is shown in Figure 1.

**Figure 1.**
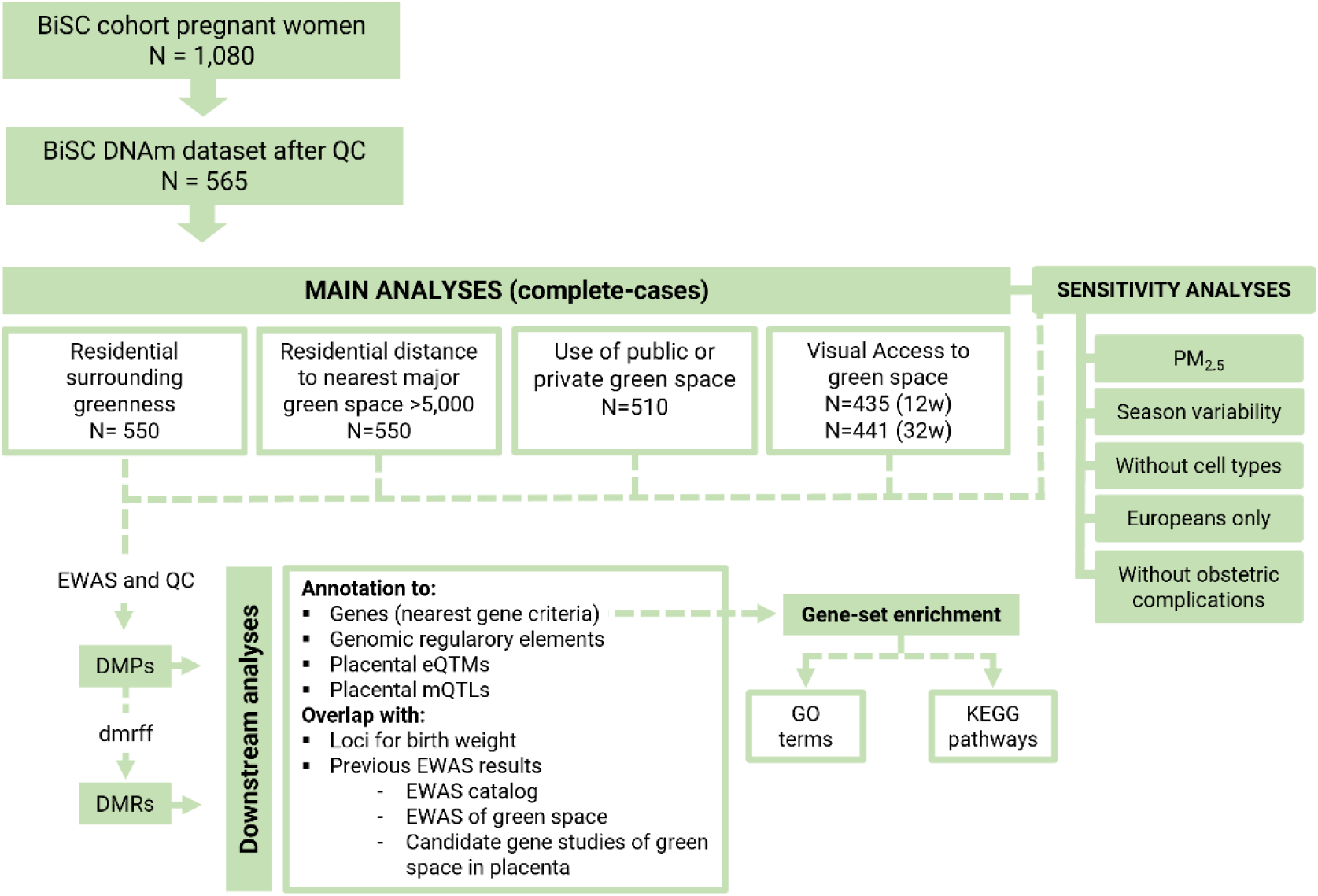
Analysis scheme. 12w: week 12 of pregnancy 32w; week 32 of pregnancy; CpG: cytosine-guanine dinucleotide; DMPs: differentially methylated positions; DMRs: differentially methylated regions; eQTM: Expression quantitative methylated regions; EWAS: Epigenome-wide association study; GO: Gene Ontology terms; KEGG: Kyoto Encyclopedia of Genes and Genomes; mQTLs: methylation quantitative trait loci; QC: quality control; PM_2.5_: particulate matter with an aerodynamic diameter <2.5 µg/m^3^; PMD: partially methylated domains.

### 2.2. Indicators of exposure to green space

We characterized four aspects of exposure to green space: (i) residential surrounding greenness, (ii) residential distance to nearest major green space as a proxy for access to green spaces, iii) use of green space, and iv) visual access to green space through the home windows.

First, to characterize residential surrounding greenness, Normalized Difference Vegetation Index (NDVI) based on high-resolution (1m x 1m) aerial images (2020) prepared by Cartographic and Geology Institute of Catalonia was used (Cartographic and Geologic Institute of Catalonia, 2021). NDVI was obtained as a ratio between the red (R) and near infrared (NIR) values in traditional fashion: (NIR-R) / (NIR + R). Its values range between −1 and 1, with higher numbers indicating more greenness (Tucker, 1979). For each participant, residential surrounding greenness was assessed as the time-weighted average NDVI in buffers of 100m, 300m and 500m during pregnancy (Gascon et al., 2016). For those participants who moved home during pregnancy, we calculated an average of these values for all residential addresses weighted by the time spent in each address during pregnancy.

Second, residential distance to nearest major green space (>5,000m^2^) was evaluated using the 2018 land use dataset from the Cartographic and Geology Institute of Catalonia (ICGC), which is based on 1m resolution imagery (Cartographic and Geologic Institute of Catalonia, 2018). The land use categories included urban, agricultural and natural green spaces.

Third, the use of green space referred to the amount of time (in hours per week) spent in green spaces during the free time in pregnancy. This indicator was self-reported and collected using questionnaires. Information about the first trimester was collected at week 12, while data on the second and third trimesters was collected at week 32 of pregnancy. The question was: “*In a typical week, during your current pregnancy, on average, how many hours of your free time do you spend in the following green spaces? (i) public parks, (ii) forest and other natural green spaces, and (iii) private garden (at home)”.* To calculate the average number of hours per week that women spent in green spaces during pregnancy, we first summed the hours spent per week in various green spaces for each trimester, including public parks, forests, other natural green spaces, and private gardens at home. Then, we computed the average across the three trimesters. A minimum of data from one trimester was required.

Fourth, visual access to green space through the home windows was self-reported and collected using questionnaire at enrolment (week 12 of pregnancy). If they changed homes, mothers were asked to respond this question again at week 32. Given the difficulty of averaging two categorical variables, we examined week 12 (first trimester) and week 32 (third trimester) separately. The question was: “*How much greenery (trees, grass, flowers, etc) can you see through the living room window? 1 = No greenery/no window; 2 = A quarter; 3 = Half; 4 = Three quarters; 5 = all is green”.* We dichotomized the answers in: 1 and 2 as visual access to greenery in less than half of the view from the living room, and 3, 4 and 5 as visual access to greenery in equal or more than half of the view from the living room window.

### 2.3. Placental biopsy and DNA extraction

Out of the 1,080 mother-child pairs that were followed until birth, 611 placentas were collected based on maternal consent and the feasibility of collection at the hospital. Biopsies were obtained by trained gynaecologists following a harmonized protocol across hospitals. Briefly, placenta biopsies of around 2.5 cm (from the maternal to the foetal side) and 1 cm width were obtained from two opposite quadrants at a distance of around 3-4 cm from site of cord insertion. Then, these biopsies were cut in two, giving a total of 4 biopsies of 2.5 x 0.5 cm. Two of them (one from each quadrant) were directly frozen in liquid nitrogen and transferred to −80C. The other two biopsies were treated with RNAlater and sent to the laboratory where they were divided in four pieces of 0.5 x 0.5 cm, corresponding roughly to the foetal membranes, the upper foetal villi, the lower foetal villi and the maternal decidua. Finally, all the biopsies were stored at −80°C for future use. Before DNA extraction, placenta samples were completely randomized and the distribution of main design and biological variables was checked among batches. For genomic DNA extraction, a fragment of approximately 5-6 cm3 (30-40 mg) was dissected from the foetal villi biopsy below the foetal membranes collected in RNAlater. All the dissection process was done in liquid nitrogen to avoid that the tissue was thawed. Then, the tissue was disrupted/homogenized using a bead mill (bead beater) at 4C for 26 seconds. Genomic DNA was then isolated using the AllPrep®DNA/RNA/miRNA Universal Kit, (Qiagen, CA, USA). DNA was eluted in 80 ul and stored in different aliquots at −80C. DNA quality was evaluated on a NanoDrop spectophotomer (Thermo Scientific, Waltham, MA, USA) and additionally 500 ng of DNA was run on 1% agarose gels to confirm that samples did not present visual signs of degradation.

### 2.4. Methylation data acquisition, quality control and normalization

DNAm assessed in 624 placental DNA samples (including 35 duplicates – 589 unique individuals) with the Infinium MethylationEPIC BeadChip from Illumina, following manufacturer’s protocol in the Human Genome facility (HUGE-F) at the Erasmus Medical Centre core facility. Briefly, 750 ng of DNA were bisulfite-converted using the EZ 96-DNAm kit following the manufacturer’s standard protocol, and DNAm measured using the Infinium protocol.

The methylation data was pre-processed using the PACEAnalysis R package (v.0.1.9) (https://www.epicenteredresearch.com/). The pre-processing pipeline consists of probe quality control, sample quality control, normalization, batch correction, winsorization of outlier values and estimation of cell type proportions. Detection p-values were estimated using out-of-band array hybridization as implemented in the SeSAMe R package (Zhou et al., 2018). Probe values were masked if the intensity values were zero, estimated based on less than three beads, and/or if they had a detection p-value>0.05. Based on these criteria (probe call rate <95%), we flagged probes that failed among at least 5% of samples for removal prior to downstream analysis (N= 63,158). Then, fifty-nine samples corresponding to 24 individuals were discarded according to: low methylated and unmethylated signal intensities overall (N=5), sample call rate < 95% (N=3), sex inconsistencies (N=9), samples with indication of substantial contamination with maternal DNA or with DNA from another participant in the study (N=11) (Heiss & Just, 2018), duplicates identified by clustering samples based on their genetic similarity using the single nucleotide polymorphisms (SNPs) included in the array (N=29) and siblings (N=2). Exclusion of one of the duplicate pairs was done randomly. A total of 565 samples remained after the sample quality control.

After removal of problematic samples, we performed pre-processing of the remaining arrays. Signal intensities were pre-processed by performing linear dye bias correction followed by single-sample background correction based on Normal-exponential convolution using out-of-band Infinium I probes (ssNoob) (J.-P. Fortin et al., 2017; Triche et al., 2013). Unwanted between-array variation was minimized by applying functional normalization using the control probes (J. P. Fortin et al., 2014). Beta-mixture quantile (BMIQ) normalization was then used to correct for the bias of Type-2 probe values (Teschendorff et al., 2013). After that, we explored the clustering of the data through Principal Component Analysis (PCA) and tested the association of the 12 first PCs with main variables (hospital, sex, ethnic origin, birth before or during SARS-CoV-2 pandemic, maternal education and maternal smoking during pregnancy) and technical variables (plate, array, extraction batch, time to placental storage, DNA concentration, and 260/280 and 260/230 ratios) to identify main batch variables. Subsequently, array batch effect was controlled with the ComBat method with the sva Bioconductor R package (Johnson et al., 2007). Following these pre-processing steps, we removed the probes flagged as problematic among the study population. Finally, to correct for the possible outliers, we winsorized the extreme values to the 1% percentile (0.5% in each side), where percentiles were estimated with the empirical beta-distribution. DNAm values are expressed as beta values, where 0 means un-methylation and 1 complete methylation.

Cell type proportions of six main placenta populations (trophoblasts, syncytiotrophoblast, nucleated red blood cell, Hofbauer cells, endothelial cells, and stromal cells) were estimated from DNAm using the reference panel from term placentas implemented in the planet R package (V. Yuan et al., 2021).

### 2.5. Covariates

The following covariates were selected *a priori* and included in the models: maternal age at recruitment (years), maternal education (no university/university), annual average household income at census tract level as a proxy of neighbourhood socioeconomic status (SES) (euros), child ethnicity (European/Latino-American/other), maternal smoking during pregnancy (no/yes), child sex (female/male), gestational age at birth (weeks), parity (nulliparous/ multiparous), hospital of birth, COVID19 confinement, which indicates whether any part of the pregnancy took place during the confinement and was categorized in three levels: pre-confinement (the whole pregnancy took place before confinement / confinement (part of the pregnancy took place during confinement) and post-confinement (the whole pregnancy took place after confinement), DNA contamination score and placental cell type proportions. Data on maternal age, maternal education, child ethnicity, maternal smoking during pregnancy and parity were self-reported and collected using a questionnaire at enrolment (week 12 of pregnancy). Annual average household income at census tract level obtained from the 2020 Standard of Living and Living Conditions survey conducted by the Spanish National Institute of Statistics (INE), which is based on fiscal data including wages, pensions, unemployment benefits, other benefits, and other income (Spanish National Institute of Statistics, 2023). Child sex was retrieved from clinical records. Gestational age at birth was calculated from the date of the last menstrual period using estimates based on the first ultrasound examination (about 12th week of gestation). DNA contamination score was calculated as the average log odds from the SNP posterior probabilities form the outlier component; capturing how irregular the SNP beta-values deviate from the ideal trimodal distribution (Heiss & Just, 2018).

### 2.6. Statistical analyses

#### 2.6.1. Differentially methylated positions (DMPs)

Epigenome-wide association studies (EWAS) using robust linear regression models were fitted to evaluate the association between maternal exposure to green space during pregnancy (predictor) and placental DNAm (outcome) using the PACEanalysis R package (https://github.com/epicenteredresearch/PACEanalysis). Main models were adjusted for the covariates described above.

We performed the quality control of the EWAS results for each model using the EASIER R package (*ISGlobal-Brge/EASIER: Tools for Methylation Data Analysis*, 2022) (Table S2). This included examining inflation and the distribution of effect estimates, standard errors and p-values. We excluded control probes, non-CpG probes, probes that mapped to X/Y chromosomes, probes with poor base pairing quality (lower than 40 on 0-60 scale), probes with non-unique 30bp 3’-subsequence (with cross-hybridizing problems), Infinium II probes with SNPs of global minor allele frequency (MAF) over 1% affecting the extension base, probes with a SNP in the extension base that causes a color channel switch from the official annotation (Zhou et al., 2017) and probes that have shown to be unreliable in a recent comparison of the Illumina 450K and EPIC BeadChips (Fernandez-Jimenez et al., 2019). The percentage of probes removed was 13.98% (Appendix A: Table S2).

Continuous green space indicators (residential surrounding greenness, residential distance to nearest major green space and use of green space) were standardized by dividing them by its interquartile range (IQR). Effect sizes are reported as the difference in DNA m unit by IQR of continuous exposures, or by exposure group of the categorical variables. Multiple testing was accounted for by applying Bonferroni correction (BN) for 694,380 tests (P-value <7.2×10^−8^). Significance was considered suggestive for p-values <1×10^−5^.

#### 2.6.2. Differentially methylated regions (DMRs)

DMRs were identified using the *dmrff* R package (Suderman et al., 2018). This method identifies candidate DMRs by screening the EWAS results for genomic regions each covered by a sequence of CpG sites with effects in the same direction, p-values <0.05, and <500 bp gaps between consecutive CpG sites. We considered statistically significant DMRs those detected with a BN p-value <0.05 and with a minimum of three consecutive CpGs within the DMR.

#### 2.6.3. Sensitivity analyses

Five sensitivity analyses were conducted by repeating the DMP and DMR analyses: i) additionally adjusting for the average exposure to particulate matter with an aerodynamic diameter <2.5 5 µg/m^3^ (PM_2.5_) as a proxy of air pollution during pregnancy ii) additionally adjusting for season of conception to investigate seasonal variability; iii) without adjusting for cell type proportions; iv) restricted to European ethnicity only, which was the largest ethnic subgroup, and v) excluding obstetric complications (preterm births, foetus with growth restriction and pregnancy complications related to the placenta: pre-eclampsia/eclampsia, gestational hypertension, placental abruption, placenta previa, oligohydramnios, polyhydramnios and chorioamnionitis).

We assessed maternal exposure to particulate matter with an aerodynamic diameter < 2.5 μm (PM_2.5_) using land use regression (LUR) models using the environmental data around collected in the BISC campaigns, following the guidelines established by the European Study of Cohorts for Air Pollution Effects (ESCAPE) (Dadvand et al., 2024). The measurement calculated around the addresses of mothers’ homes was used to adjust the model of the residential green space indicators (surrounding greenness, distance and visual access), while the measurement considering all environments (home, workplace, and commuting route) was used to adjust the model of use of green space.

Season of conception (autumn / spring / summer / winter) was calculated by subtracting the gestational days of pregnancy (last menstrual period) from the date of birth. Preterm was defined as a gestational age at birth of less than 37 weeks. Foetal growth restriction was defined as a birth weight percentile <3 or a birth weight percentile <10 and cerebroplacental ratio pulsatility index percentile at 32w <5 or a birth weight percentile <10 and uterine artery pulsatility index percentile at 32w >95. Birth weight percentiles were estimated according to the BCNatal curves (Figueras et al., 2008; Figueras & Gratacós, 2014). Pulsatility indexes were measured through Doppler and percentiles calculated using reference curves described elsewhere (Baschat & Gembruch, 2003; Gómez et al., 2008).

### 2.7. Downstream analyses

First, we tested the overlap of the BN significant DMPs and DMRs with placental chromatin states (Ernst & Kellis, 2017), placenta germline differently methylated regions (gDMRs) (Hamada et al., 2016), and placenta partially methylated domains (PMDs) (Schroeder et al., 2013). Second, to assess whether the methylation levels of the DMP and DMRs were associated with the expression levels of nearby genes, we examined previously identified Expression Quantitative Trait Methylation (eQTMs) in the placenta (Delahaye et al., 2018; Deyssenroth et al., 2020). Third, we checked whether the BN significant DMPs and CpGs within the significant DMRs had previously been associated with exposures or health traits using the information from the EWAS Catalog (Battram, Yousefi, et al., 2022). (Battram, Yousefi, et al., 2022). Fourth, we compared the list of BN significant DMPs and DMRs with previously reported studies investigating the association between exposure to green space during pregnancy and DNAm in cord blood (Aguilar-Lacasaña et al., 2024; Alfano et al., 2023) and a candidate gene study in placenta (Dockx et al., 2022).

As DNAm is partially regulated by genetic variants, we explored whether the DMP and CpGs in the DMRs found in our study had any methylation Quantitative Trait Loci (mQTLs). Foetal cis mQTLs (+-0.5 Mb, 1 Mb window) were identified in the BiSC study (n=408) by applying a linear regression for each CpG-SNP pair adjusting for child’s sex, child’s gestational age at birth, child’s ancestry based on 5 genetic principal components (PCs), 6 placental cell type proportions estimated from DNA methylation data using the TensorQTL tool (Ongen et al., 2016). Genome-wide significance was stablished at 5×10-8. Placental DNAm was measured with the EPIC array as described above, and foetal genome-wide genotyping (placenta or cord blood) was conducted using the Infinium Global Screening (GSA) array v.3.0 array and imputed with the Haplotype Reference Consortium (HRC) panel.

Additionally, we assessed the overlap with genomic regions of previously identified SNPs for birth weight in the largest genome-wide association study (GWAS) to date (Juliusdottir et al., 2021). The overlap was investigated using the the GenomicRanges R package (Lawrence et al., 2013) and by defining 1 Mb windows (+-0.5 Mb) surrounding each of the autosomal SNPs identified in the GWAS.

Then, the DMP and DMRs were annotated to genes using the Illumina’s annotation file, which annotates CpGs located within a distance of 1500bp from the transcription start site (TSS). If the CpG did not have a gene annotated within this distance, then the University of California, Santa Cruz (UCSC) Genome Browser was referenced to identify the nearest gene using the matchGenes() function from the bumphunter R package (Jaffe et al., 2012). We conducted functional enrichment analyses for Gene Ontology (GO) terms (Kanehisa, 2000) and pathways of the Kyoto Encyclopedia of Genes and Genomes (KEGG) (Kanehisa, 2000) of genes annotated to the suggestive DMPs (p-value <1×10^−5^) and CpGs within the BN significant DMRs, using the missMethyl method (Phipson et al., 2016) as implemented in EASIER R package (ISGlobal-Brge/EASIER: Tools for Methylation Data Analysis, 2022).

## 3. Results

### 3.1. Study population

A total of 550 participants were included in the study. A detailed description of our study population characteristics is presented in Table 1; Appendix A: Table S1. The median age of the mothers was 34.4 years old, around two thirds had university education and 7.0% smoked during pregnancy. The median gestational age at birth was 40.0 weeks, half of the offspring were girls, 67.5% of them were Europeans, 28.9% Latino-American, and the rest from other ethnicities. Median residential NDVI value was 0.2, the median residential distance to a green space was 129.9 m and a median of 3 h per week were spent in public and/or private green spaces. Furthermore, around one-third of the mothers had visual access to greenery in more than half of the living room window during the first and third trimesters. The correlation coefficients across the different indicators of residential surrounding greenness (NDVI 100m, 300m and 500m) were high, especially between the 300m and 500m buffers. For visual access, the correlation between the first and third trimesters was also very high. We found inverse moderate correlations across residential surrounding greenness and residential distance to a major green space. The correlation coefficients across all green space exposures are shown in Appendix B: Figure S1.

**Table 1.**
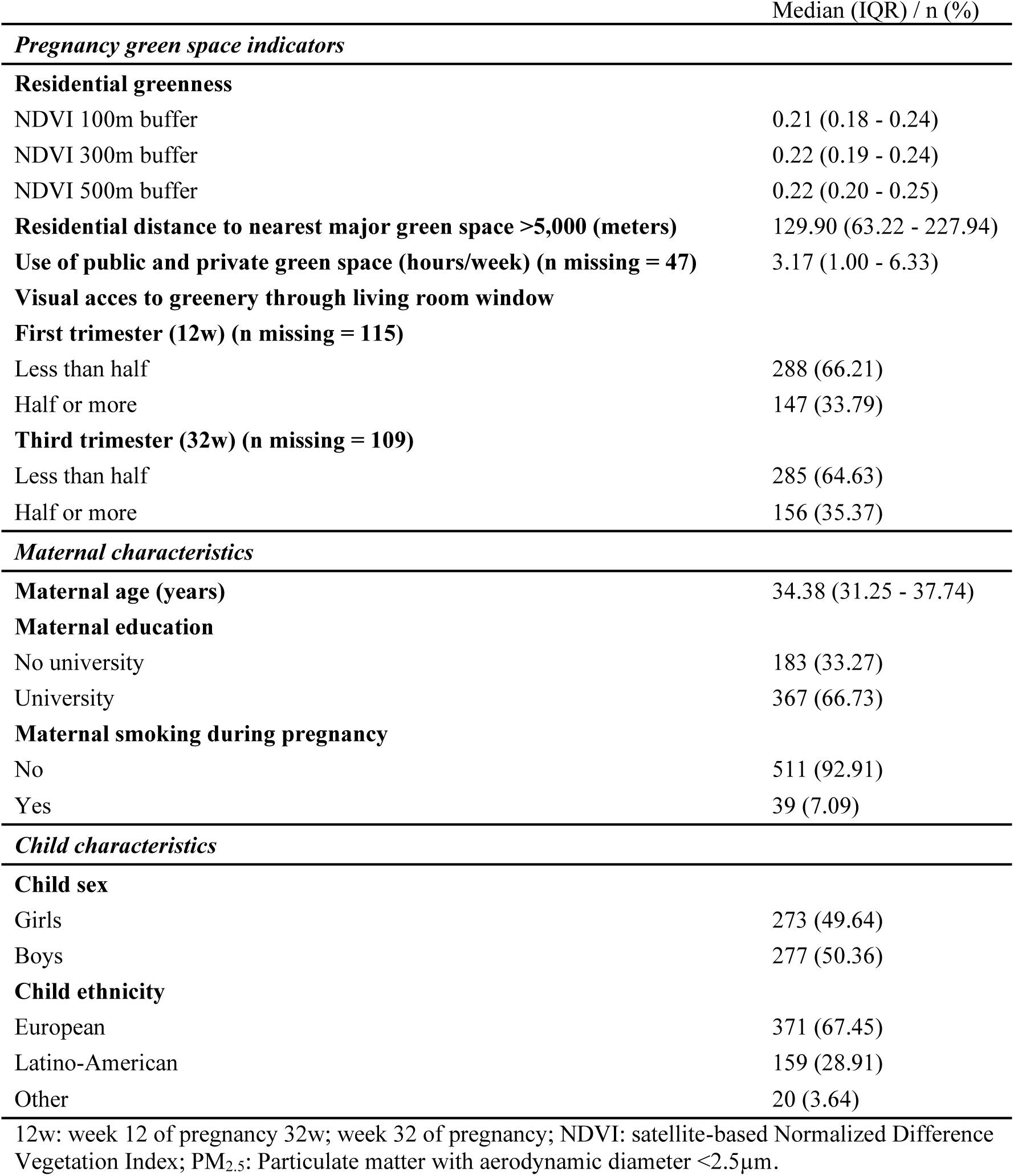
Characteristics of the BiSC study population (N= 550)<colcnt=1>.

### 3.2. Green space during pregnancy and placental DNAm

#### 3.2.1. DMPs

The lambda inflation factors for the main models ranged from 0.84 to 1.26 (Appendix A: Table S2; Appendix B: Figure S2). After BN correction, one DMP (cg14852540) annotated to the *SLC25A10* gene was significantly associated with residential surrounding greenness in 500m buffer. At this DMP, for an IQR increase of NDVI in 500m, DNAm was 0.3% lower. The effect at this site remained consistent in both the 100m and 300m buffers, although not statistically significant (Appendix A: Table S3). No DMPs were significantly associated with residential distance, use or visual access to green space. At suggestive significance (p-value <1×10^−5^), 101 unique DMPs were associated with at least one green space indicator (Figure 2A; Appendix A: Table S3). Pearson correlation coefficients of the effect estimates of all genome-wide CpGs and the BN DMP across the EWAS results from the green space indicators are shown in Appendix A: Table S4; Appendix B: Figure S3.

**Fig 2.**
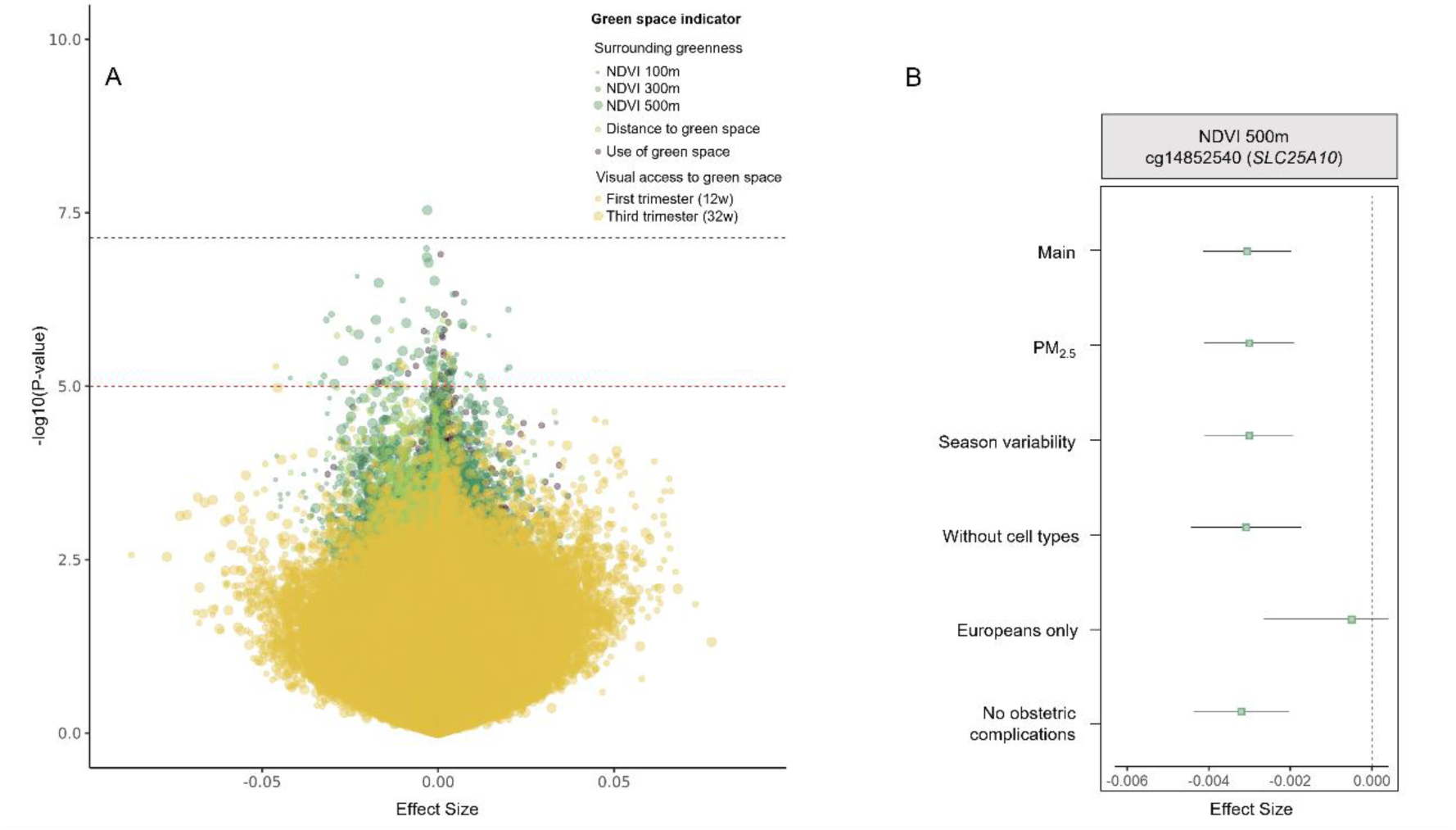
(A) Volcano plot showing the effect sizes on the x-axis and the (-log_10_) p-values on the y-axis for the association between all pregnancy green space indicators and placental DNAm. Each dot represents the association of one CpG with one exposure variable. The color of the dot indicates the exposure: Surrounding greenness (dark green), distance to green space (light green), use of green space (purple) and visual access to green space (yellow). Dot sizes correspond to the sizes of the NDVI buffers and time window of visual access (week 12 and week 32 of pregnancy). The black line represents the BN p-value threshold (7.2×10^−8^) and the red line represents the suggestive p-value (1×10^−5^). (B) Forest plot showing the effect size and 95% confidence interval of the BN significant DMP in the main model compared with the sensitivity analyses: 1) adjusted for PM_2.5_ during pregnancy; 2) adjusted for season of conception; 3) without adjusting for cell type proportions; 4) restricted to European ethnicity, and 5) without obstetric complications (preterm births, cases of FGR and pregnancy complications).

In terms of sensitivity analyses, the most notable difference in effect size among the BN significant DMP was observed when we restricted the analyses to Europeans (n=371), leading to an 83.6% decrease in the effect estimate (Figure 2B). Effect sizes did not change substantially in the rest of sensitivity analyses. The percentage differences in absolute values in the effects between the main and the sensitivity results were as follows: a decrease of 1.6% for PM_2.5_ adjustment, a decrease of 1.5% for season of conception adjustment, an increase of 0.6% for cell type proportion unadjusted model and an increase of 5.5% when excluding obstetric complications (Figure 2B; Appendix A: Table S5).

#### 3.2.2. DMRs

Ten unique DMRs, annotated to 10 genes, were associated with pregnancy exposure to different green space indicators after BN correction. Residential surrounding greenness in 500m was associated with four DMRs, two of which were also associated with 300m buffer. Distance to green space was associated to five DMRs and visual access to green space during the first trimester was inversely associated with one DMR. This DMR, associated with visual access during the first trimester, was no longer identified with this exposure in the third trimester. However, an overlapping DMR was detected with nominal significance (p-value = 2.77×10^−5^) (Appendix A: Table S6). These DMRs included from three to eight CpGs with widths ranging from 20 to 382 bp. No DMPs were significantly associated with residential surrounding greenness in 100m, use of green space or visual access to green space during the third trimester. More detailed information on each DMR is provided in Table 2. Additionally, the genes annotated to the significant DMRs that overlap across the evaluated green space indicators are shown in Figure 3.

**Fig. 3.**
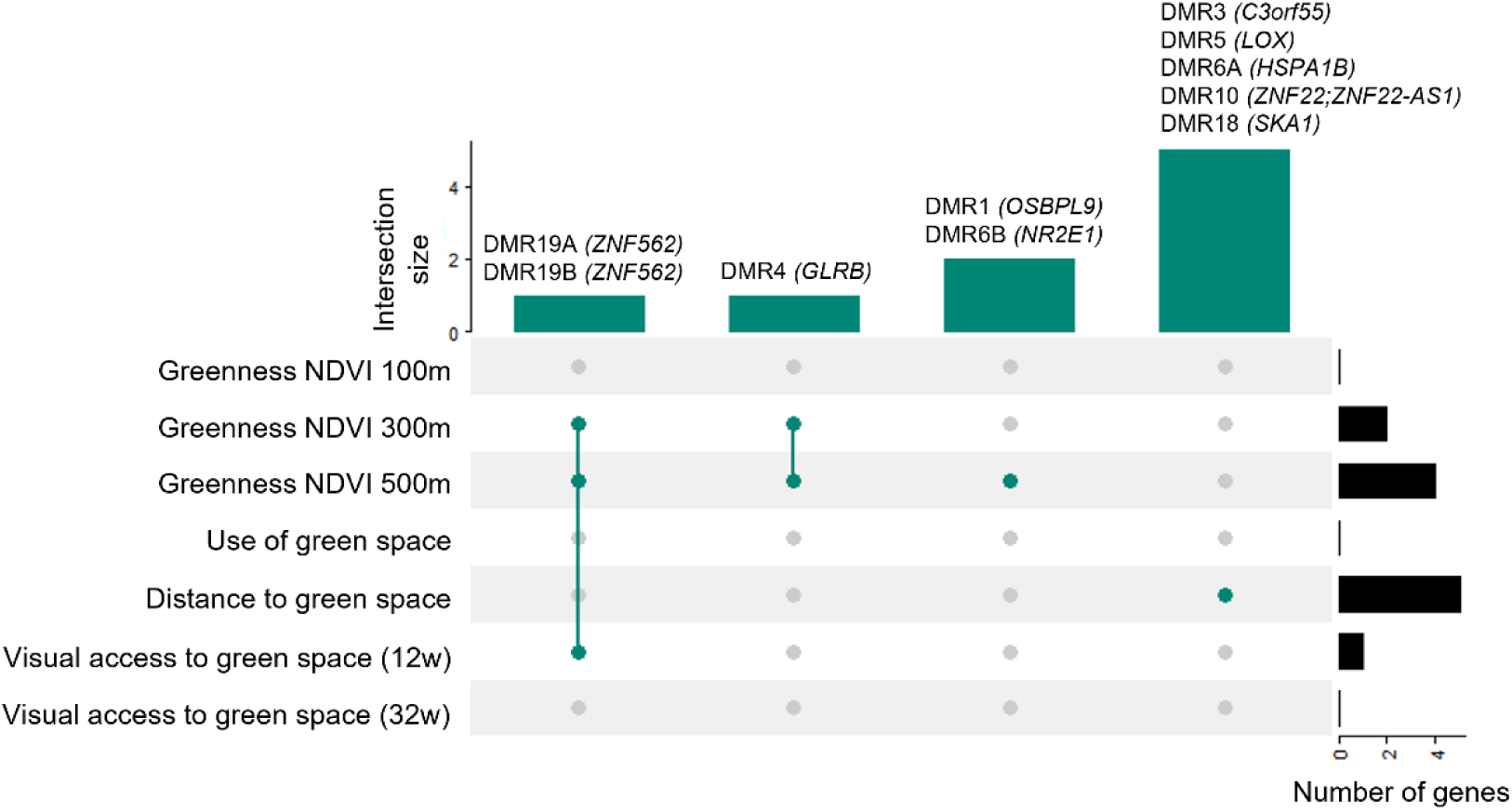
Upset plot representing the complete or partial overlap of significant DMRs across the different green space indicators. The dots indicate the presence (green) or absence (grey) of DMRs overlapping across the exposure categories. The green bars indicate the number of DMRs that overlap across the green space indicators. The black bars on the right indicate the total number of genes annotated to a DMR for each green space indicator. 12w: week 12 of pregnancy; 32w: week 32 of pregnancy.

**Table 2.**
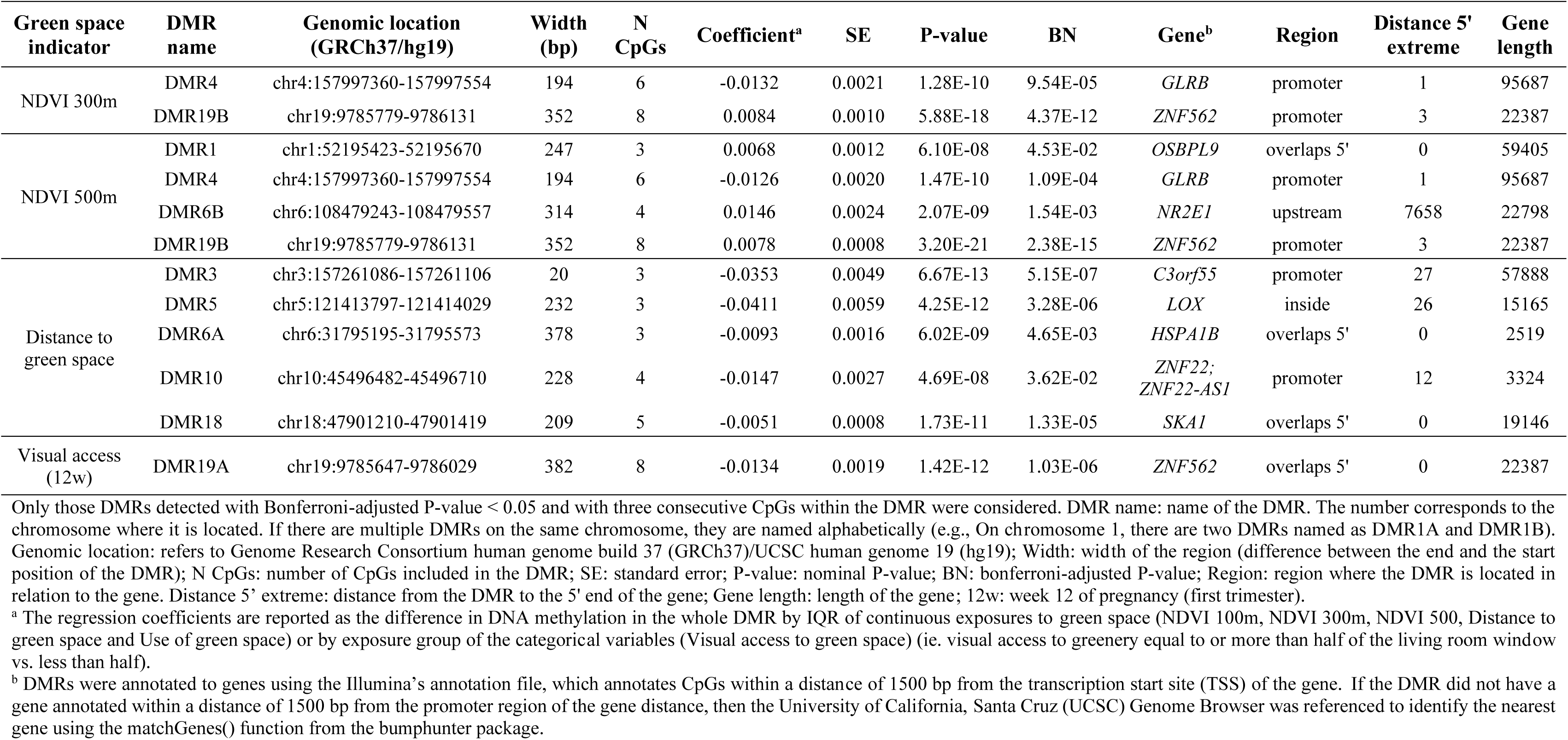
Differentially methylated regions (DMRs) associated with green space indicators in placenta.

| Green space indicator | DMR name | Genomic location (GRCh37/hg19) | Width (bp) | N CpGs | Coefficient<sup>a</sup> | SE | P-value | BN | Gene<sup>b</sup> | Region | Distance 5' extreme | Gene length |
| --- | --- | --- | --- | --- | --- | --- | --- | --- | --- | --- | --- | --- |
| NDVI 300m | DMR4 | chr4:157997360-157997554 | 194 | 6 | -0.0132 | 0.0021 | 1.28E-10 | 9.54E-05 | *GLRB* | promoter | 1 | 95687 |
|  | DMR19B | chr19:9785779-9786131 | 352 | 8 | 0.0084 | 0.0010 | 5.88E-18 | 4.37E-12 | *ZNF562* | promoter | 3 | 22387 |
| NDVI 500m | DMR1 | chr1:52195423-52195670 | 247 | 3 | 0.0068 | 0.0012 | 6.10E-08 | 4.53E-02 | *OSBPL9* | overlaps 5' | 0 | 59405 |
|  | DMR4 | chr4:157997360-157997554 | 194 | 6 | -0.0126 | 0.0020 | 1.47E-10 | 1.09E-04 | *GLRB* | promoter | 1 | 95687 |
|  | DMR6B | chr6:108479243-108479557 | 314 | 4 | 0.0146 | 0.0024 | 2.07E-09 | 1.54E-03 | *NR2E1* | upstream | 7658 | 22798 |
|  | DMR19B | chr19:9785779-9786131 | 352 | 8 | 0.0078 | 0.0008 | 3.20E-21 | 2.38E-15 | *ZNF562* | promoter | 3 | 22387 |
| Distance to green space | DMR3 | chr3:157261086-157261106 | 20 | 3 | -0.0353 | 0.0049 | 6.67E-13 | 5.15E-07 | *C3orf55* | promoter | 27 | 57888 |
|  | DMR5 | chr5:121413797-121414029 | 232 | 3 | -0.0411 | 0.0059 | 4.25E-12 | 3.28E-06 | *LOX* | inside | 26 | 15165 |
|  | DMR6A | chr6:31795195-31795573 | 378 | 3 | -0.0093 | 0.0016 | 6.02E-09 | 4.65E-03 | *HSPA1B* | overlaps 5' | 0 | 2519 |
|  | DMR10 | chr10:45496482-45496710 | 228 | 4 | -0.0147 | 0.0027 | 4.69E-08 | 3.62E-02 | *ZNF22;*<br>*ZNF22-AS1* | promoter | 12 | 3324 |
|  | DMR18 | chr18:47901210-47901419 | 209 | 5 | -0.0051 | 0.0008 | 1.73E-11 | 1.33E-05 | *SKA1* | overlaps 5' | 0 | 19146 |
| Visual access (12w) | DMR19A | chr19:9785647-9786029 | 382 | 8 | -0.0134 | 0.0019 | 1.42E-12 | 1.03E-06 | *ZNF562* | overlaps 5' | 0 | 22387 |

Only those DMRs detected with Bonferroni-adjusted P-value < 0.05 and with three consecutive CpGs within the DMR were considered. DMR name: name of the DMR. The number corresponds to the chromosome where it is located. If there are multiple DMRs on the same chromosome, they are named alphabetically (e.g., On chromosome 1, there are two DMRs named as DMR1A and DMR1B). Genomic location: refers to Genome Research Consortium human genome build 37 (GRCh37)/UCSC human genome 19 (hg19); Width: width of the region (difference between the end and the start position of the DMR); N CpGs: number of CpGs included in the DMR; SE: standard error; P-value: nominal P-value; BN: bonferroni-adjusted P-value; Region: region where the DMR is located in relation to the gene. Distance 5' extreme: distance from the DMR to the 5' end of the gene; Gene length: length of the gene; 12w: week 12 of pregnancy (first trimester).
<sup>a</sup> The regression coefficients are reported as the difference in DNA methylation in the whole DMR by IQR of continuous exposures to green space (NDVI 100m, NDVI 300m, NDVI 500, Distance to green space and Use of green space) or by exposure group of the categorical variables (Visual access to green space) (ie. visual access to greenery equal to or more than half of the living room window vs. less than half).
<sup>b</sup> DMRs were annotated to genes using the Illumina's annotation file, which annotates CpGs within a distance of 1500 bp from the transcription start site (TSS) of the gene. If the DMR did not have a gene annotated within a distance of 1500 bp from the promoter region of the gene distance, then the University of California, Santa Cruz (UCSC) Genome Browser was referenced to identify the nearest gene using the matchGenes() function from the bumpHunter package.

In the sensitivity analyses, the smallest difference in the number of significant DMRs was observed after adjusting for the season of conception, with 9 out of 10 DMRs remaining significant. In contrast, the largest difference in the number of significant DMRs was seen when restricting the analysis to Europeans only, detecting 4 out of the 10 significant DMRs from the main model. The absolute percentage differences in effect across the main and sensitivity results for DMRs that remained significant varied as follows: 0.4% to 2.67% for PM_2.5_ adjustment, 0.8% to 8.1% for season of conception adjustment, 0.3% to 24.1% for unadjusted cellular composition, 11.3% to 22.6% when restricted to Europeans only, and 1.7% to 14.2% for the model excluding obstetric complications (Appendix A: Table S7).

#### 3.3.3. Downstream analyses

We searched whether the BN significant DMP and DMRs (42 CpG sites in total) were located in specific genomic regulatory elements of the placenta. More than 80% (34/42) were located in active regions: active TSS (TssA), flanking active TSS (TssAFlink) and enhancers (Enh). Moreover, DMR18 (chr18:47901210-47901419) overlapped with two placental PMDs, regions known to contain relevant genes for placental function (Schroeder et al., 2013). However, none of the DMP or DMRs overlapped with placental gDMRs (Appendix A: Table S8). Regarding gene expression, none of the significant DMP or DMRs overlapped with the eQTMs identified in the placenta (Appendix A: Table S8). According to genetic variation, the DMP (cg14852540) and one DMR (DMR3) were close to two fetal SNPs associated with birth weight (rs73354194 and rs13322435) (Juliusdottir et al., 2021). However, neither of these CpGs has a cis-mQTL in placenta (Appendix A: Table S8).

As one BN significant DMP was not enough to perform a gene-set enrichment analyses, we included those DMPs associated to a green space indicator at the suggestive threshold (p-value <1×10^−5^) and CpGs within the BN DMRs. Although not significant after multiple testing correction, among the top pathways where these genes were involved, glucocorticoid synthesis and secretion (cortisol), inflammatory response, regulation of response to reactive oxygen species, and longevity regulating pathways, were among the top ones (nominal p-value <0.1) (Appendix A: Table S9-S10).

Neither the DMP nor the DMRs found in this study overlapped with previous findings or vice versa when evaluating the association between green space exposure and DNAm in cord blood (Aguilar-Lacasaña et al., 2024; Alfano et al., 2023) or in placenta (Dockx et al., 2022) (Appendix X: Table S11).

According to the EWAS Catalog, methylation levels at the significant DMP or CpGs within the significant DMRs have previously been related to child age, gestational age, autoimmune diseases such as rheumatoid arthritis and respiratory conditions such as chronic obstructive pulmonary disease (COPD). None of these studies were conducted in placenta and most of them evaluated in blood (see Appendix A: Table S8 for detailed information).

## 4. Discussion

This study evaluated association of different aspects of maternal exposure to green space during pregnancy and placental DNAm in the BiSC cohort. We found that this exposure was associated with one DMP (cg14852540 annotated to *SLC25A10* gene) and 10 unique DMRs.

The DMP that we identified in relation to residential greenness within 500m buffer was annotated to the gene body of the *Solute carrier family 25 member 10* (*SLC25A10*). *SLC25A10*, a dicarboxylate carrier, plays a role in metabolic processes by transporting small molecules into or out of the mitochondria thereby providing substrates for metabolic processes including the Krebs cycle and fatty acid synthesis (Freund et al., 2014; Huypens et al., 2011; Mizuarai et al., 2005). Buccal mitochondria DNA content (mtDNAc), a proxy of mitochondrial function (Castellani et al., 2020), has been positively associated with green space exposure (Hautekiet et al., 2022). Furthermore, mitochondrial respiration has been shown to play a role in the invasive and migratory capabilities of trophoblasts, which are necessary for embryo implantation and placental development (Xiong et al., 2024; Yu et al., 2024). Additionally, recent studies found that DNAm of this gene in blood was associated with body mass index and waist circumference in adolescents (Huang et al., 2022). Finally, this DMP was located at <0.5 Mb of a loci containing foetal SNPs associated with birth weight (Juliusdottir et al., 2021), which may suggest the involvement of this genomic region in foetal growth.

Besides this DMP, we also identified 10 DMRs. Two of them (DMR19A and DMR19B) were associated with residential greenness and visual access to green space, respectively. These DMRs partially overlapped in the promoter region of *the Zinc Finger Protein 562* (*ZNF562*) gene. Despite being in the same region, these DMRs showed opposite effects. The reason for these differences is unclear, and further investigation is needed. Moreover, we observe that DMR19B, which was associated with visual access during the first trimester, did not remain significant after multiple testing in the third trimester. However, an overlapping DMR was detected with nominal significance, and with the same direction of effect. This gene may be involved in transcriptional regulation, and a recent study suggests that it might be related to glutamine metabolism its involvement in glutamine metabolism (Shi et al., 2024), an amino-acid essential for foetal growth, especially in later gestation (Wu et al., 2015). In a previous study, a placental DMR (chr19:9785647-9785919) covering this gene and overlapping our DMRs was linked to a “vegetarian tendency” pattern (Lecorguillé et al., 2022).

Additionally, we found that residential surrounding greenness was associated with two other DMRs (DMR1, DMR4) annotated to *OSBPL9* and *GLRB*, respectively. First, DMR1 overlapped the 5’ untranslated region (5’UTR) of *the Oxysterol Binding Protein Like 9 (OSBPL9)* gene, known for its role in lipid metabolism (Paterson et al., 2022). Second, DMR4 located in the promoter of the *Glycine Receptor Beta (GLRB)* gene was inversely associated with residential surrounding greenness within 300m and 500m buffer. *GLRB* encodes the beta subunit of glycine receptors, which are neurotransmitter-gated ion channels found throughout the central nervous system, including the hippocampus, spinal cord, and brain stem (Handford et al., 1996). This gene has been associated with neurological disorders such as hyperekplexia (L. Zhou et al., 2002).

Distance to green space was also associated to DMRs located inside or near the promoter of the following genes: *Solute Carrier Family 66 Member 1 Like, Pseudogene* (*C3orf55*) participating in L-lysine transport and close to a SNP associated with birth weight (Juliusdottir et al., 2021); *Lysile Oxidase* (*LOX*) involved in collagen and elastin cross-linking (Guo et al., 2016)*; Spindle And Kinetochore Associated Complex Subunit 1 (SKA1*), crucial for proper chromosome segregation (Welburn et al., 2009); *Heat Shock Protein Family A (Hsp70) Member 1B (HSPA1B)*, playing a role in placenta-derived stem cells in the heat-induced proteotoxic stress response, which is crucial for protecting the proteome against stress (Alharbi et al., 2022); and Zinc Finger Protein family genes such as *Zinc Finger Protein 22* (*ZNF22*) and *ZNF22 Antisense RNA 1 (ZNF22-AS1),* may be involved in transcriptional regulation (Paterson et al., 2022), although limited findings in relation to these genes have been published so far.

Among the top gene-sets and GO and KEGG terms identified, although not surviving multiple-testing, there were glucocorticoid related pathways, longevity regulating pathway, inflammatory response, and regulation of response to reactive oxygen species. The genes in the glucocorticoid-related pathways whose DNAm levels were suggestively associated with green spaces in our study were *Adenylate Cyclase 4 (ADCY4), Aryl Hydrocarbon Receptor Interacting Protein (AIP)* and *Phosphodiesterase 8A (PDE8A)* genes. Glucocorticoids have a central role in foetal maturation, but excessive levels can induce foetal growth restriction (Fowden et al., 2016). According to the literature, stress reduction could be one of the mechanisms linking green exposure to improved health outcomes through epigenetics (Nwanaji-Enwerem et al., 2024). Moreover, in a previous study, higher residential surrounding greenness, residential proximity to green spaces, longer time spent in green spaces, and more visual access to green space during pregnancy were associated with lower cortisol level in cord blood (Boll et al., 2020). Therefore, larger studies with increased statistical power are needed to explore this pathway concerning green space and its potential implications for placental and foetal development.

To date, two recent studies evaluated the association between residential greenness exposure during the prenatal period, a critical period for perinatal and life-long health, with DNAm in cord blood (Aguilar-Lacasaña et al., 2024; Alfano et al., 2023). None of the DMP or DMRs identified in this study overlapped with the findings from the aforementioned studies. This could be explained by the differences between tissues (Ohgane et al., 2008), but also differences in the exposure and outcome assessment.

Alfano et al., used land cover data to classify green space based on vegetation height, rather than NDVI, to calculate green space exposure. Additionally, the buffer of 100m was the only one common to both studies. In our analysis, we did not observe any

significant DMP or DMR within this buffer. Moreover, the lack of overlap with our previous study (Aguilar-Lacasaña et al.,) may be due to the different resolutions of the aerial images used, with the current study having a much higher resolution (1m x 1m) compared to the previous study (30 x 30m).

In placenta, only a candidate gene study (N=327) has explored the association between green space and DNAm (Dockx et al., 2022). In that study, methylation levels at the promoter of *5-Hydroxytryptamine Receptor 2A (HTR2A)* gene, a serotonin receptor with a potential effect on neurodevelopment function, was positively associated with maternal green space exposure within 1,000 m, 2,000 m and 3,000 m buffers measured using land cover data and stratified into low (<3 m) and high (≥3 m) vegetation. We were unable to assess the same CpGs in our study as they were not included in the EPIC array. Therefore, we examined the direction of the effect of nearby CpGs located in the promoter region (cg27068143 and cg10323433) of *HTR2A*, and although not statistically significant (nominal P-value >0.05), they showed a consistent direction of the effect in our study.

One of the strengths of this study was the analysis of a relatively large number of placental samples, a relevant tissue for foetal development. Second, to characterize residential surrounding greenness, NDVI was derived from high-resolution (1×1) aerial images and averaged over the entire pregnancy, considering the changes in participants’ residences. Moreover, we expanded our analyses beyond surrounding greenness by including other green space indicators such as distance to nearest major green space, use of green spaces and also visual access to greenery through the home window. Third, DMRs were detected using the *dmrff* R package, considered the gold standard for calculating DMRs (Lent et al., 2021). Fourth, relying on wealth of available data in BiSC, we were able to conducted sensitivity analyses by further adjusting our analyses for air pollution, season of conception, cellular composition or excluding pregnancy and foetal complications, and we observed that our findings were generally robust after conducting these analyses.

It is important to interpret the results of this study considering its limitations. First, despite being the largest study on placenta evaluating the association of prenatal exposure to green space and DNAm, our study might have been underpowered for detecting some of the associations. For example, the decrease in the number of significant DMRs observed when restricting the analysis to European ethnicity (N= 371), compared to the main analysis including all ethnicities, may be attributed to the decrease in statistical power due to the reduction in sample size. Also, the effect size of the significant DMP was reduced substantially in the subset of Europeans, suggesting some residual confounding by ethnic origin not well controlled in the full population. We note that models were adjusted for ethnic origin. Larger sample sizes are needed to investigate ethnicity and ancestry specific effects. Moreover, we did not examine sex-specific or cell-interacting associations due to statistical power limitations. Second, we did not have data in other important aspects of the green space exposure, such as type of vegetation and quality characteristics of the green space. Third, the variable measuring the use of green space was obtained from questionnaire, which could have resulted in exposure misclassification. Validating this measure by employing other techniques such as mobile phone Global Positioning System (GPS) data would improve its reliability (Heikinheimo et al., 2020). Fourth, whilst we controlled our analyses for a wide array of covariates, we cannot assume that associations we found are causal. In the same way, we cannot assume that differences in DNAm will affect health outcomes, as epigenetic mechanisms are more complex that what can be discerned from DNAm (Min et al., 2021). Finally, in addition to being unable to explore associations with other epigenetic marks and gene expression, this study, in common with other EWAS studies, covers only a small proportion of the epigenome (less than 5% of the 23 million CpGs for the EPIC array) (Battram et al., 2022).

## 5. Conclusion

Overall, we identified associations between green space exposure during pregnancy and DNAm levels in placenta at one DMP and ten DMRs annotated to genes involved in transcriptional regulation, metabolic pathways and mitochondrial respiration. This points to a potential role of placental epigenetic mechanisms in the effects of green space exposure during pregnancy on birth outcomes and offspring health. However, further research is needed to validate these results and understand the underlying biological pathways.

## Data Availability

Genome wide DNA methylation summarized results can be found at Zenodo repository (THIS WILL BE DONE UPON ACCEPTANCE OF THE MANUSCRIPT). Individual cohort level data may be available by application to the relevant institution after obtaining required approvals.

## Abbreviations

ADCY4: Adenylate Cyclase 4 gene
AIP: Aryl Hydrocarbon Receptor Interacting Protein
BiSC: Barcelona Life Study Cohort
BMIQ: Beta-mixture quantile
BN: Bonferroni
Bp: base pairs
C3orf55: Solute Carrier Family 66 Member 1 Like, Pseudogene
CpG: Cytosine-phosphate-guanine
COPD: chronic obstructive pulmonary disease
DMP: Differentially methylated position
DMR: Differentially methylated region
DNAm: DNA methylation
IQR: Interquartile range
eQTM: Expression quantitative methylated regions
ESCAPE: European Study of Cohorts for Air Pollution Effects
Enh: Enhancer
EWAS: Epigenome-wide association study
gDMR: Germline differently methylated region
GLRB: Glycine Receptor Beta gene
GPS: Global Positioning System
GS: Green space
GO: Gene Ontology
HSPA1B: Heat Shock Protein Family A (Hsp70) Member 1B gene
HTR2A: 5-Hydroxytryptamine Receptor 2A gene
HUGE-F: Human Genome facility
ICGC: Cartographic and Geology Institute of Catalonia
KEGG: Kyoto Encyclopedia of Genes and Genomes
LOX: Lysile Oxidase gene
LUR: Land Use Regression Models
mQTL: methylation Quantitative Trait Loci
mtDNAc: mitochondria DNA content
NR2E1: Nuclear Receptor Subfamily 2 Group E Member 1 2
OSBPL9: Oxysterol Binding Protein Like 9 gene
PCA: Principal Component Analysis
PDE8A: Phosphodiesterase 8A gene
PM2.5: Particulate matter with aerodynamic diameter 2.5 microm
PMD: Partially methylated domain
QC: Quality control
R: Red values
MAF: minor allele frequency
NIR: near infrared values
NDVI: Normalized Difference Vegetation Index
SKA1: Spindle And Kinetochore Associated Complex Subunit 1 gene
SLC25A10: Solute carrier family 25 member 10 gene
SNPs: single nucleotide polymorphisms
SES: socioeconomic status
TSS: transcription start site
TssA: active transcription start site
TssAFlink: flanking active transcription start site
UCSC: University of California, Santa Cruz
UTR: untranslated region
ZNF22: Zinc Finger Protein 22
ZNF22-AS1: Zinc Finger Protein 22 Antisense RNA 1
ZNF562: Zinc Finger Protein 562 gene.

## CRediT authorship contribution statement

**Sofía Aguilar-Lacasaña:** Conceptualization, Methodology, Formal analysis, Investigation, Resources, Data curation, Writing - Original Draft, Writing - Review & Editing, Visualization. **Marta Cosin-Tomas:** Conceptualization, Methodology, Resources, Writing-review & editing, Supervision. **Bruno Raimbault:** Data curation, Methodology, Writing - Review & Editing. **Laura Gómez:** Resources, Data curation, Writing - Review & Editing. **Olga Sánchez:** Resources, Writing-review & editing. **Maria Julia Zanini:** Resources, Writing-review & editing. **Rosalia Pascal Capdevila:** Resources, Writing-review & editing. **Maria Foraster:** Resources, Writing-review & editing. **Mireia Gascon:** Resources, Writing-review & editing. **Ioar Rivas:** Resources, Writing-review & editing. **Elisa Llurba:** Resources, Writing-review & editing. **Maria Dolores Gómez-Roig:** Resources, Writing-review & editing. **Jordi Sunyer:** Conceptualization, Investigation, Writing - Review & Editing, Supervision, Funding acquisition. **Mariona Bustamante:** Conceptualization, Methodology, Investigation, Writing - Review & Editing, Supervision, Funding acquisition. **Martine Vrijheid:** Conceptualization, Investigation, Writing - Review & Editing, Supervision, Funding acquisition. **Payam Dadvand:** Conceptualization, Methodology, Investigation, Writing - Review & Editing, Supervision, Funding acquisition.

## Acknowledgments

This work was supported by the European Joint Programming Initiative “A Healthy Diet for a Healthy Life” (JPI HDHL and Instituto de Salud Carlos III) – NutriPROGRAM (AC18/00006), Instituto de Salud Carlos III and co-funded by European Union (ERDF) “A way to make Europe” – ALMA project (PI20/00190) and the European Union’s Horizon 2020 research and innovation program – ATHLETE project (874583). The BiSC cohort was supported by the European Research Council (ERC) under the European Union’s Horizon 2020 research and innovation programme – AirNB project (785994) and the Health Effects Institute (HEI), an organization jointly funded by the United States Environmental Protection Agency (EPA) (R-82811201) and certain motor vehicle and engine manufacturers. We would like to thank all the participants and their families for their generous collaboration. A full list of BiSC researchers can be found at https://projectebisc.org/en/team/. Genome-wide genotyping data was funded by the Instituto de Salud Carlos III (ISCIII) and co-funded by European Union (ERDF) “A way to make Europe” – ENTENTE project (PI20/01116) and the Centro Nacional de Genotipado-CEGEN (PRB2-ISCIII). Sofía Aguilar-Lacasaña was funded by the European Union’s Horizon 2020 research and innovation program – ATHLETE project (874583) and the FI-AGAUR Predoctoral contract [2023 FI-2 00797]. Marta Cosin-Tomas was funded by a Beatriu de Pinós Postdoctoral Contract awarded by Generalitat de Catalunya-AGAUR and European Commission-Horizon 2020 (2019 BP 00107). Mireia Gascon holds a Miguel Servet fellowship (Grant CP19/00183) funded by Acción Estratégica de Salud - Instituto de Salud Carlos III, co-funded by European Social Fund “Investing in your future”. We acknowledge support from the grant CEX2023-0001290-S funded by MCIN/AEI/ 10.13039/501100011033, and support from the Generalitat de Catalunya through the CERCA Program.

## Ethics approval and consent to participate

From pregnancy up to 18 months’ visits were approved by the Clinical Research Ethics Committee of the Parc de Salut Mar project (2018/8050/I), Medical Research Committee of the Fundació de Gestió Sanitària del Hospital de la Santa Creu i Sant Pau de Barcelona (EC/18/206/5272), and Ethics Committee of the Fundació Sant Joan de Déu (PIC-27-18). Before joining the cohort during their regular first trimester hospital visit, participants were informed by a BiSC midwife or nurse about the study’s details, duration, and their option to withdraw without penalty. If they agreed to take part, they signed consent forms permitting the collection of biological samples and genetic studies, receiving a copy for themselves. Parents or legal guardians of children also provided informed consent before any study procedures commenced. Specific consent forms for genetic studies were also provided to mothers and parents or legal guardians of children.

## Consent for publication

Not applicable.

## Data availability

Genome-wide DNA methylation summarized results can be found at Zenodo repository (https://zenodo.org/) [THIS WILL BE DONE UPON ACCEPTANCE OF THE MANUSCRIPT]. Individual cohort level data may be available by application to the relevant institution after obtaining required approvals. The code for the analyses is available in this GitHub repository link: https://github.com/sofiaguilarl/EWAS_GreenSpace_Placenta.git.

## Appendix A. Supplementary tables

## Appendix B. Supplementary figures

