## Appendix B: Supplementary figures for "Epigenome-wide association study of pregnancy exposure to green space and placental DNA methylation"

### **Figure S1.** Correlation plot across green space exposure indicators during pregnancy. The correlation coefficient across the green space indicators was estimated with the Spearman’s correlation for continuous variables, and with the polyserial correlation for the combination of categorical and continuous variables and polychoric correlation for the combination of two categorical variables.

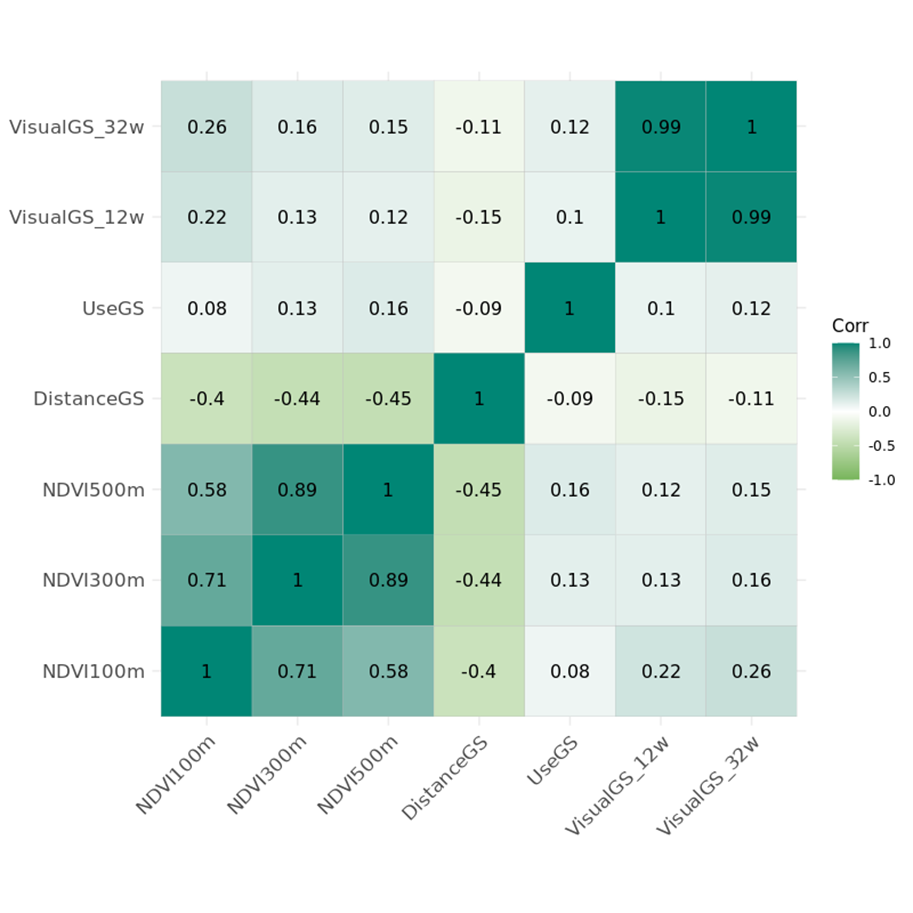

### **Figure S2.** QQ-plots and genomic inflation factors (lambdas) of the model adjusted for cell type proportions (main model) of green space during pregnancy and placental DNA methylation: (A) NDVI 100m, (B) NDVI 300m, (C) NDVI 500m, (D) Distance to green space, (E) Use of green space, (F) Visual access to green space (12w), (G) Visual access to green space (32w).

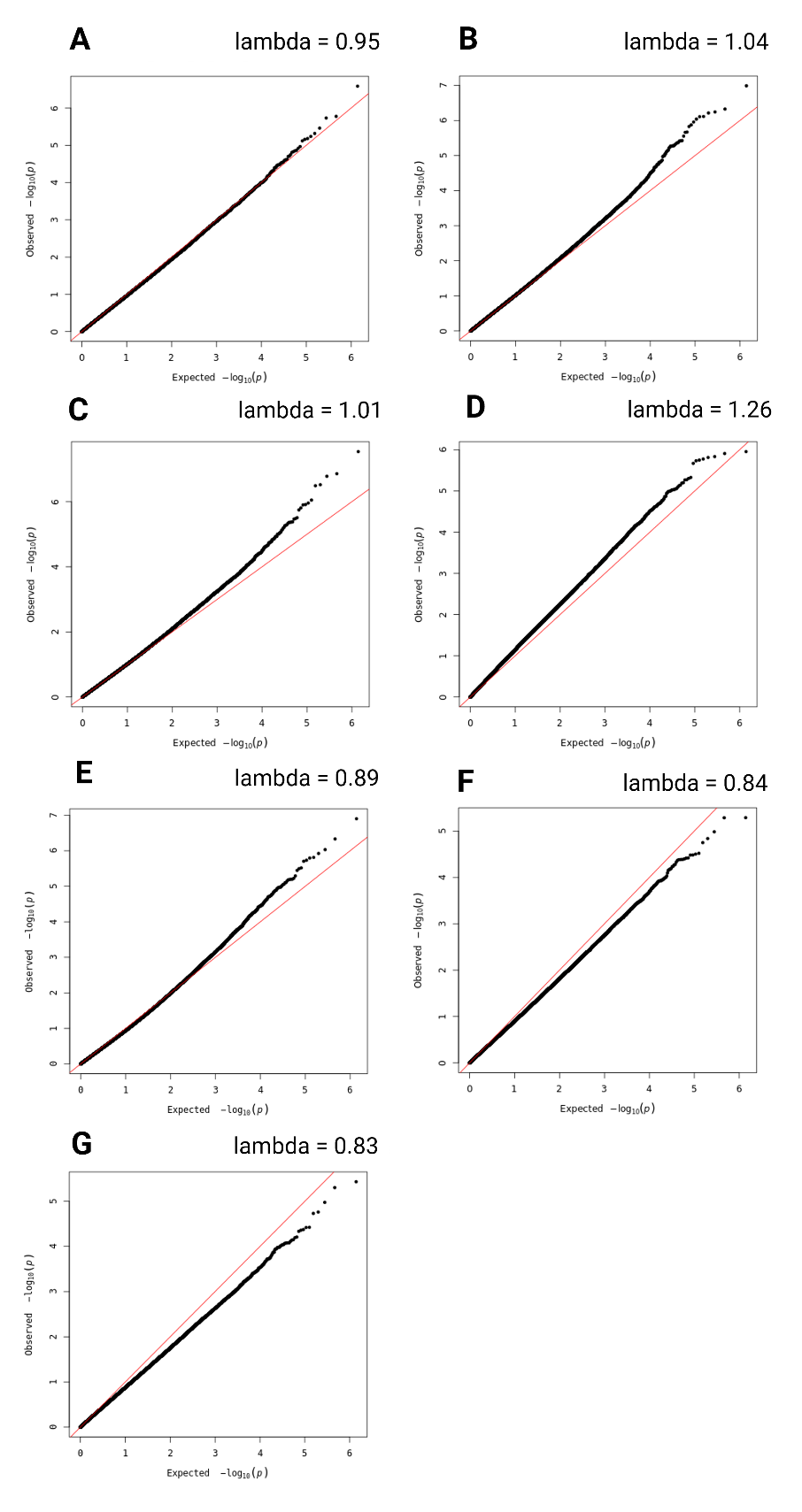

### **Figure S3**. Correlation plot of the effect size of all genome-wide CpGs across green space indicators during pregnancy.

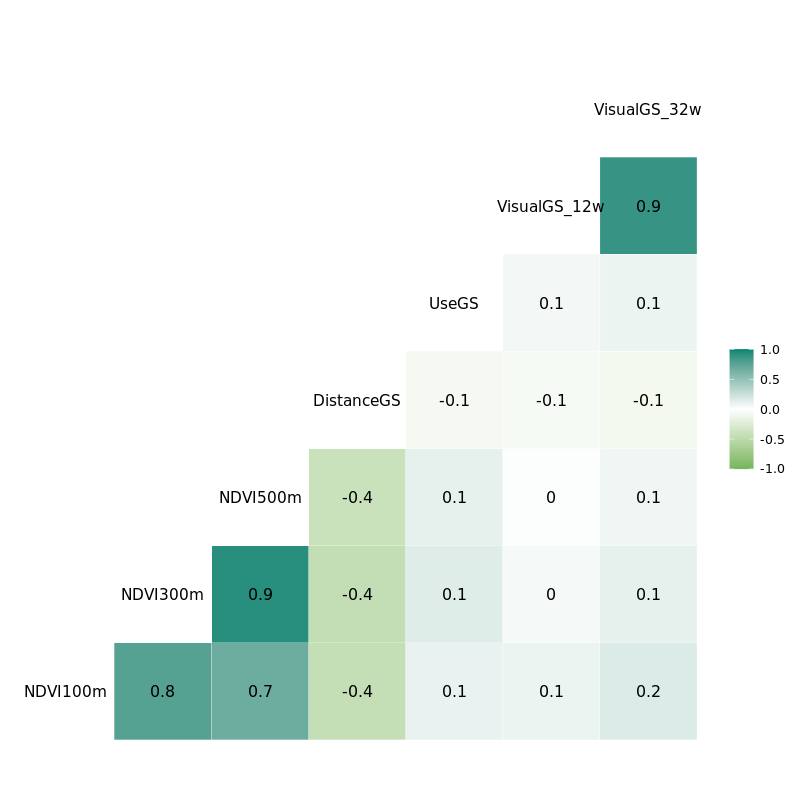
